# Analysing the psychosocial and health impacts of Long COVID in Pakistan: A cross sectional study

**DOI:** 10.1101/2023.05.22.23290323

**Authors:** Madeeha Khan, Sadaf Majeed, Qura Tul Ain, Amjad Nawaz, Khadija Awais Sumra, Vilma Lammi, Faizan Nihal, Aleena Afrah, Ejaz Ahmed Khan, Mohammad Iqbal Khan, Fouzia Sadiq

## Abstract

Long COVID corresponds to the occurrence of symptoms beyond twelve weeks after the onset of acute COVID-19 illness. The study aimed to analyze impacts of long COVID on the general health and psychosocial well-being of the Pakistani population. This cross-sectional study aimed to analyse the impacts of long COVID on general health and psychosocial well-being. For this study, the participants were interviewed, and their responses were recorded on a questionnaire capturing information on demographics, COVID-19 status, duration of symptoms and long COVID symptoms. The psychological impacts of the pandemic were assessed using scales like Short Mood and feeling questionnaire (sMFQ), Warwick-Edinburgh Mental Well-being Scale (WEMWBS), Generalized Anxiety Disorder Assessment (GAD-7) and Perceived Stress Scale (PSS). Regression analysis was conducted to analyse the predictors of long COVID. A total of 300 participants were interviewed, of which 155 (52%) had COVID-19 illness. Of these 54 (35%) had persistent symptoms for a period of more than 12 weeks classified as long COVID. Muscle problems and fatigue were the most frequent (14.7%) symptoms encountered, followed by breathing problems (12.6%) and cognitive issues (12.6%). Symptoms such as decrease in appetite and confusion or disorientation during the initial phase of the infection were associated with long COVID. Majority of the participants (83.3%) had moderate level of perceived stress while moderate to severe levels of stress was observed in 17.3% of the individuals. Moreover, a high level positive mental wellbeing was also observed.

## Introduction

Since the onset of COVID-19 pandemic in 2020, over 600 million cases have been reported resulting in a loss of more than 6 million lives (*WHO Coronavirus (COVID-19) Dashboard | WHO Coronavirus (COVID-19) Dashboard With Vaccination Data*, 2023.). In Pakistan, more than 1.57 million cases and 30,629 deaths by COVID-19 have been reported so far (*COVID-19 Health Advisory Platform by Ministry of National Health Services Regulations and Coordination*, 2023.). The acute phase of COVID-19 presents with varied clinical manifestations, ranging from respiratory, musculoskeletal, and gastrointestinal to neurological symptoms (Proal & VanElzakker, 2021). A significant proportion of COVID-19 survivors present with persistent symptoms even after weeks or months of the infection, collectively known as long-COVID or post COVID condition/syndrome (Proal & VanElzakker, 2021). Long COVID corresponds to occurrence of symptoms in individuals with a history of probable or confirmed SARS-CoV-2 infection, usually three months after the onset of COVID-19, that cannot be explained by alternative diagnosis (Lopez-Leon et al., 2021). Most commonly reported symptoms include fatigue, headache, dyspnea, attention disorder, and hair loss; however, several other respiratory, cardiovascular, and neurocognitive symptoms have also been reported (Lopez-Leon et al., 2021)]. These symptoms can be continued or persistent symptoms of acute infection, relapsing symptoms, or new symptoms without any alternative diagnosis (Raveendran et al., 2021). The prevalence of long COVID symptoms is influenced by many factors such as severity of the acute infection, age, and sex (Raveendran et al., 2021).

Several pathophysiological pathways have been identified for the pathogenesis of long COVID including: 1) direct viral tissue damage caused by the high binding ability of SARS-CoV-2 spike protein to the angiotensin-converting enzyme 2 (ACE2) at a variety of locations in human body, facilitating virus entry to target cells 2) endothelial cell damage that can trigger excessive thrombin production in the vascular beds leading to thrombosis 3) dysregulation of immune system and inflammatory response which can lead to autoimmune processes or post-acute SARS-CoV-2 infection (Gupta et al., 2020)]. Moreover, prolonged viral shedding, particularly in immunocompromised patients, and psychological factors such as post-traumatic stress disorder can also contribute to the long-term persistence of COVID-19 symptoms (Raveendran et al., 2021),(Aydillo et al., 2020). Neuro-oxidative mechanisms, such as increased oxidative toxicity and lowered antioxidant defenses may also mediate post viral somatic and mental symptoms (Al-Hakeim et al., 2022).

Besides deteriorating health issues, long COVID also poses detrimental impacts on the healthcare system with regards to providing long-term support and services to these patients (Munblit et al., 2022). Moreover, it can be a contributing factor for socioeconomic burden, as the affected individuals may face long-term consequences such as longer breaks from work and working for reduced hours, leading to unemployment and subsequently financial hardships (Nittas et al., 2022).

The present study aims to evaluate the symptoms, and the influence of different factors on the development of long COVID in the Pakistani subjects. This study also aims to explore the impacts of COVID-19 pandemic on the psychological health and social activities of the study participants.

## Methods

### Study design and participants

A cross sectional study was conducted across Pakistan. The study was approved by Institutional Review Board & Ethics Committee (IRB&EC), Shifa Tameer-e-Millat University Islamabad (IRB # 274-21). The questionnaire was adapted from the “COVID APP” (*ZOE Health Study*, 2022) and “The Avon Longitudinal Study of Parents and Children (ALSPAC COVID)” (Smith et al., 2021) questionnaires and amended according to the context of Pakistan. A convenience sampling approach was employed, and the participants were interviewed during September 2021 to February 2022. Informed consent and were taken from the participants prior to the interview. The data analysis was completed by September 2022.

### Procedures

The questionnaire was composed of questions based on i) general questions i.e., age, ethnicity, gender, residence, and blood group ii) general health before the pandemic iii) COVID disease status i.e., confirmation of COVID-19, symptoms, recovery, hospitalization, and duration of the disease iii) long COVID status; persistent or onset of new symptoms beyond 12 weeks of COVID-19 disease iv) difficulties and support for long COVID patients v) psychosocial impacts of COVID-19 pandemic (Supplementary material).

The participants of the study were characterized as: Long-COVID (those who experienced one or more symptoms beyond 12 weeks or three months after acute phase of COVID-19 illness), Normal COVID (those with acute infection but recovered within 12 weeks), No COVID (healthy participants who either did not contract the illness or were asymptomatic).

### Analysis of psychosocial impacts of COVID-19 pandemic

To assess the impacts of COVID-19 pandemic on mental well-being, several well-known scales were incorporated in the study, including short Mood and feeling questionnaire (sMFQ), Warwick Edinburgh Mental Wellbeing Scale (WEMWBS), Generalized Anxiety Disorder Assessment (GAD-7), and Perceived Stress Scale (PSS) (Angold et al., 1995),(Tennant et al., 2007),(Löwe et al., 2008),(Cohen et al., 1983)]. The details of the instruments and the questions are provided in the Supplementary material.

### Statistical analysis

The data was collected, analysed, and visualized in Microsoft Office 365 Excel (Microsoft Corporation, USA) and IBM SPSS Statistics, version 26.0 (IBM, USA). The distribution of continuous variables was presented as mean ± standard deviation (Std) or as median and interquartile range (IQR), whereas categorical data was presented as frequencies and percentages. Multinomial logistic regression model was performed to assess the association of gender, age, ethnicity, blood group, and smoking status with dependent variable (no covid, long covid, normal covid), where no COVID was taken as reference category and Long COVID, normal COVID were taken as comparative categories. Binary logistic regression model was performed to assess the association of independent variables (demographic features and symptoms) with the binary response of outcome variable (long COVID and normal COVID). Odds ratio (OR) and 95% confidence interval (95% CI) of each variable were calculated. The forest plot for regression analysis were created using Python v3.11.0. Mann-Whitney U test and Kruskal Wallis H test was used to assess the psychological impacts of COVID-19 pandemic among healthy, normal COVID and long COVID group.

## Results

### Characteristics of the study participants

A total of 300 respondents were included in the study and final analysis. The mean age of the participants was 32±12.4 years. Among these, 200 (66.7%) were female, while 100 (33.3%) were male. Majority of the participants were from Punjab (n=148; 49.3%), followed by Islamabad Capital territory (n=130; 43.3%), Khyber Pakhtunkhwa (n=13; 4.3%), Sindh (n=7; 2.3%), Balochistan (n=1; 0.3%), and Azad Kashmir (n=1; 0.3%). 155 (51.7%) had COVID-19 either confirmed by positive RT-PCR and/or diagnosed by a general physician. The dates for the infection ranged from March 2020 to November 2021. The demographic characteristics of the study participants are given in Table 1.

**Table 1.** Demographic details of the study participants

| **Characteristics of the respondents** | **Overall**<br>**(n=300)** | **No COVID**<br>**(n=145)** | **Normal COVID**<br>**(n=101)** | **Long COVID**<br>**(n=54)** |
| --- | --- | --- | --- | --- |
| **Age (Years)** |  |  |  |  |
| Mean±SD | 32±12.4 | 31.26±11.6 | 31.45±12.4 | 34.5±14.3 |
| **N (%)** |  |  |  |  |
| <20 | 33 (11) | 17 (11.7) | 10 (10.0) | 6 (11.1) |
| 20-40 | 200 (66.7) | 102 (70.3) | 70 (69.0) | 28 (52.0) |
| 40-60 | 57 (19) | 21 (14.5) | 18 (18.0) | 18 (33.3) |
| >60 | 10 (3.3) | 5 (3.4) | 3 (3.0) | 2 (3.7) |
| **Gender** |  |  |  |  |
| Male | 100 (33.3) | 51 (35.1) | 35 (34.6) | 14 (26.0) |
| Female | 200 (66.7) | 94 (64.8) | 66 (65.34) | 40 (74.0) |
| **Ethnicity** |  |  |  |  |
| Punjabi | 201 (67) | 96 (66.2) | 66 (65.3) | 39 (72.2) |
| Pathan | 42 (14) | 21 (14.4) | 16 (16.0) | 5 (9.2) |
| Urdu Speaking | 33 (11) | 18 (12.4) | 11 (11.0) | 4 (7.4) |
| Kashmiri | 8 (2.7) | 2 (1.4) | 2 (2.0) | 4 (7.4) |
| Other | 10 (3.3) | 3 (2.0) | 5 (4.6) | 2 (3.7) |
| **Smoking status** |  |  |  |  |
| Smoker | 17 (5.7) | 7 (5) | 6 (5.9) | 4 (7.4) |
| Nonsmoker | 283 (94.3) | 138(95) | 95 (94.0) | 50 (92.6) |
| **Blood group** |  |  |  |  |
| A+ | 20 (6.7) | 6 (4.1) | 9 (8.91) | 5 (9.25) |
| A- | 6 (2.0) | 3 (2.0) | 2 (1.9) | 1 (1.8) |
| B+ | 47 (15.7) | 19 (13.1) | 19 (18.8) | 9 (16.6) |
| B- | 3 (1.0) | 0 (0.0) | 3 (2.9) | 0 (0.0) |
| AB+ | 11 (3.7) | 5 (3.4) | 3 (2.9) | 3 (5.5) |
| AB- | 1 (0.3) | 0 (0.0) | 0 (0.0) | 1 (1.8) |
| O+ | 31 (10.3) | 9 (6.2) | 8 (7.9) | 14 (25.9) |
| O- | 3 (1.0) | 2 (1.4) | 1 (1) | 0 (0.0) |
| Unknown | 178 (59.3) | 101 (69.6) | 56 (55.4) | 21 (38.9) |
| **General health before COVID-19 pandemic** |  |  |  |  |
| Excellent | 89 (29.7) | 46 (31.7) | 31 (30.7) | 12 (22.2) |
| Very good | 87 (29.0) | 43(29.6) | 33 (32.6) | 11 (20.4) |
| Good | 97 (32.3) | 42 (28.9) | 32 (31.6) | 23 (42.6) |
| Fair | 26 (8.7) | 13 (8.9) | 5 (4.9) | 8 (14.8) |
| Poor | 1 (0.3) | 1 (0.7) | 0(0.0) | 0 (0.0) |

### Symptoms during COVID-19 illness

Fever and feeling feverish were the most common symptoms felt/observed during the acute phase of the illness, reported by 124 (80%) and 101 (65%) of the individuals respectively (Supplementary Figure 1). Fatigue or feeling of unusual tiredness was experienced by 98 (63%) individuals, followed by sore throat (n=96; 62%), headache, anosmia (loss or change in sense of smell; n=81; 52%), and ageusia (loss of or change in sense of taste; n=76; 49%) (Supplementary Figure 1). Medical help i.e., discussion of symptoms with a doctor or general practitioner was sought by 113 (73%) of the infected individuals, while only 13 (8.4%) were hospitalized (Supplementary Table 2).

### Long COVID Symptoms

Out of the 155 COVID-19 positive patients, 54 (35%) experienced symptoms beyond 12 weeks of infection. A small proportion (n=9; 16.7%) of these long haulers were hospitalized during the acute phase. Among these, 22.2% developed a new condition that might have association with COVID as diagnosed by their physician (Supplementary Table 3). Post viral fatigue was one of the most common newly diagnosed conditions reported by 38.1% of the participants. Most frequent symptoms, experienced beyond the 12-week period, were muscle problems and fatigue (14.7%) followed by breathing difficulties such as breathlessness, dyspnea, painful breathing, or cough (12.6%) and cognitive issues relating to communicating and thinking such as brain-fog, memory issues, difficulty concentrating, decreased alertness, confusion, difficulty speaking (12.6%). Other symptoms such as altered sense of taste and smell (anosmia or ageusia), problems relating to mood such as anxiety or feeling down/irritable, headaches were reported by 8.4% of the individuals (Figure 1).

Based on the data collection timespan of this study, and the information of the circulating variants of concern during that period (*GISAID - Gisaid.Org*, n.d.),(*COVID-19 | Institute for Health Metrics and Evaluation*, n.d.), most of the COVID-19 positive patients (n=155) might have been infected by the ancestral SARS-CoV-2 strain (n=59; 38%), followed by delta (B.1.351; n=50; 32%) and alpha (B.1.1.7; n=46; 30%) variants. Similarly, most of the long COVID patients (n=54) might have been infected by the ancestral SARS-CoV-2 strain (n=28; 52%), followed by alpha (B.1.1.7; n=16; 30%) and delta (B.1351; n=10; 18%) variant.

### Predictors of Long COVID

In multinomial regression model, age [p=0.04; OR=1.02 (95% CI=1.00-1.05)], gender [p=0.03; OR=2.44 (95%CI= 1.08-5.52)] and blood group [p=0.00; OR=0.84 (95% CI= 0.75-0.94) were the significant predictors of long COVID among the first set of covariates (Figure 2a). Blood group was the significant predictor among the second group of covariates [p=0.13; OR=0.89 (95% CI= 0.81-0.97)] compared to the healthy participants (Figure 2b). In binary regression analysis, presence of symptoms such as decrease in appetite [p=0.01; OR=0.30 (95% CI= 0.11-0.81)] and confusion/disorientation/drowsiness [p=0.05; OR=0.38 (95% CI= 0.15-1.00)] during the acute phase of the COVID were statistically significant with long COVID compared to normal COVID (Figure 3).

### Difficulties experienced during daily routine

When asked about the level of difficulties in conducting daily tasks such as taking care of household responsibilities, physical activity, social activities, and day to day work, 12 weeks beyond COVID-19 illness, the majority faced no difficulties in these tasks. Twenty four percent (n=13) faced extreme difficulties in walking for a long distance (1 km or half a mile) (Supplementary Figure 2). Family (44.3%) and friends (23.7%) were identified by the patients as their most important source of support while coping with long COVID.

### Psychosocial impacts of COVID-19 pandemic

The analysis based on sMFQ showed that depression was present (Score ≥12) among 15% (n=45) of the study participants. No significance difference was observed in groups based on gender, age, ethnicity and COVID status (Table 2). The assessment of positive mental wellbeing assessed by WEMWBS showed a higher positive metal wellbeing in the 82.7% of the participants (n=248). Among the study participants, 57% (n=171) had minimal anxiety levels, while moderate and severe anxiety was found in 10.3% (n=31) and 7% (n=21) of the individuals, respectively as determined by the GAD-7 scale. No significant differences were observed in different groups. Moderate stress levels were observed in most of the participants (83.3%, n=250) as determined by the PSS-10 scale (Table 2). The median score for sMFQ was 2 (IQR=9.00-0.00), where significant differences were observed in the median sMFQ scores (p=0.00) among the three groups (long COVID, normal COVID and no COVID). The median score of WEMWBS was 51 (IQR=12). For GAD-7, the median score was 3 (IQR=7.00-1.00) suggesting minimum anxiety in the study participants. The median score of PSS was 18 (IQR=22.00-16.00) which indicates presence of moderate stress among the study participants (Supplementary Table 3).

**Table 2:**
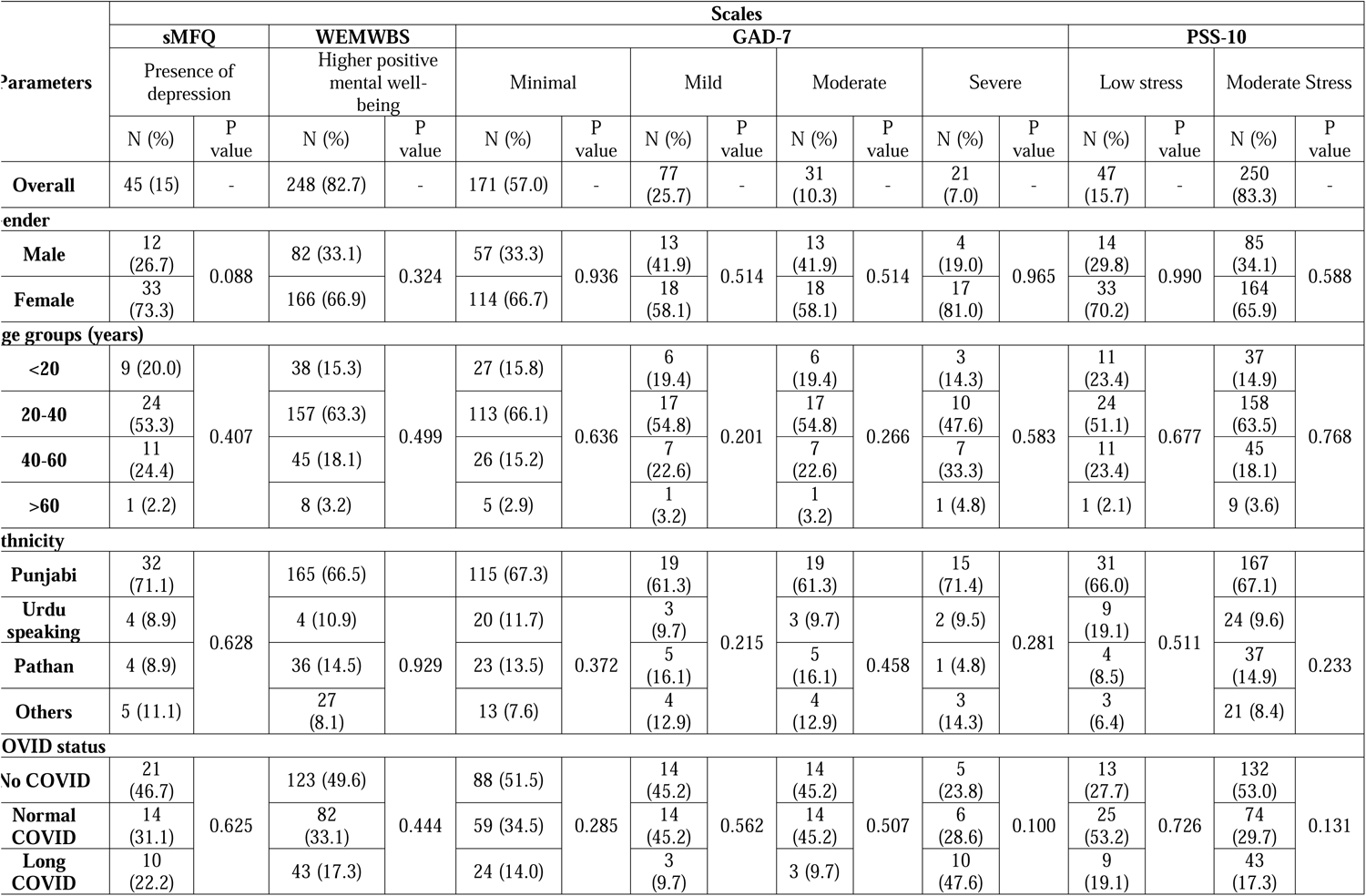
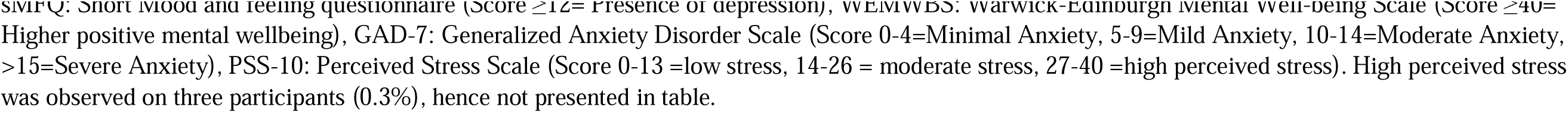
Comparison of psychological impacts of COVID-19 pandemic among different groups

| Parameters | Scales |  |  |  |  |  |  |  |  |  |  |  |  |  |  |  |
| --- | --- | --- | --- | --- | --- | --- | --- | --- | --- | --- | --- | --- | --- | --- | --- | --- |
|  | sMFQ |  | WEMWBS |  | GAD-7 |  |  |  |  |  |  |  | PSS-10 |  |  |  |
|  | Presence of depression |  | Higher positive mental well-being |  | Minimal |  | Mild |  | Moderate |  | Severe |  | Low stress |  | Moderate Stress |  |
|  | N (%) | P value | N (%) | P value | N (%) | P value | N (%) | P value | N (%) | P value | N (%) | P value | N (%) | P value | N (%) | P value |
| Overall | 45 (15) | - | 248 (82.7) | - | 171 (57.0) | - | 77 (25.7) | - | 31 (10.3) | - | 21 (7.0) | - | 47 (15.7) | - | 250 (83.3) | - |
| Gender |  |  |  |  |  |  |  |  |  |  |  |  |  |  |  |  |
| Male | 12 (26.7) | 0.088 | 82 (33.1) | 0.324 | 57 (33.3) | 0.936 | 13 (41.9) | 0.514 | 13 (41.9) | 0.514 | 4 (19.0) | 0.965 | 14 (29.8) | 0.990 | 85 (34.1) | 0.588 |
| Female | 33 (73.3) |  | 166 (66.9) |  | 114 (66.7) |  | 18 (58.1) |  | 18 (58.1) |  | 17 (81.0) |  | 33 (70.2) |  | 164 (65.9) |  |
| Age groups (years) |  |  |  |  |  |  |  |  |  |  |  |  |  |  |  |  |
| <20 | 9 (20.0) | 0.407 | 38 (15.3) | 0.499 | 27 (15.8) | 0.636 | 6 (19.4) | 0.201 | 6 (19.4) | 0.266 | 3 (14.3) | 0.583 | 11 (23.4) | 0.677 | 37 (14.9) | 0.768 |
| 20-40 | 24 (53.3) |  | 157 (63.3) |  | 113 (66.1) |  | 17 (54.8) |  | 17 (54.8) |  | 10 (47.6) |  | 24 (51.1) |  | 158 (63.5) |  |
| 40-60 | 11 (24.4) |  | 45 (18.1) |  | 26 (15.2) |  | 7 (22.6) |  | 7 (22.6) |  | 7 (33.3) |  | 11 (23.4) |  | 45 (18.1) |  |
| >60 | 1 (2.2) |  | 8 (3.2) |  | 5 (2.9) |  | 1 (3.2) |  | 1 (3.2) |  | 1 (4.8) |  | 1 (2.1) |  | 9 (3.6) |  |
| Ethnicity |  |  |  |  |  |  |  |  |  |  |  |  |  |  |  |  |
| Punjabi | 32 (71.1) | 0.628 | 165 (66.5) | 0.929 | 115 (67.3) | 0.372 | 19 (61.3) | 0.215 | 19 (61.3) | 0.458 | 15 (71.4) | 0.281 | 31 (66.0) | 0.511 | 167 (67.1) | 0.233 |
| Urdu speaking | 4 (8.9) |  | 4 (10.9) |  | 20 (11.7) |  | 3 (9.7) |  | 3 (9.7) |  | 2 (9.5) |  | 9 (19.1) |  | 24 (9.6) |  |
| Pathan | 4 (8.9) |  | 36 (14.5) |  | 23 (13.5) |  | 5 (16.1) |  | 5 (16.1) |  | 1 (4.8) |  | 4 (8.5) |  | 37 (14.9) |  |
| Others | 5 (11.1) |  | 27 (8.1) |  | 13 (7.6) |  | 4 (12.9) |  | 4 (12.9) |  | 3 (14.3) |  | 3 (6.4) |  | 21 (8.4) |  |
| COVID status |  |  |  |  |  |  |  |  |  |  |  |  |  |  |  |  |
| No COVID | 21 (46.7) | 0.625 | 123 (49.6) | 0.444 | 88 (51.5) | 0.285 | 14 (45.2) | 0.562 | 14 (45.2) | 0.507 | 5 (23.8) | 0.100 | 13 (27.7) | 0.726 | 132 (53.0) | 0.131 |
| Normal COVID | 14 (31.1) |  | 82 (33.1) |  | 59 (34.5) |  | 14 (45.2) |  | 14 (45.2) |  | 6 (28.6) |  | 25 (53.2) |  | 74 (29.7) |  |
| Long COVID | 10 (22.2) |  | 43 (17.3) |  | 24 (14.0) |  | 3 (9.7) |  | 3 (9.7) |  | 10 (47.6) |  | 9 (19.1) |  | 43 (17.3) |  |

SIMFQ: Short mood and feeling questionnaire (Score $\geq 12$ = Presence of depression), WEMWBS: warwick-Edinburgh Mental well-being Scale (Score $\geq 40$ = Higher positive mental wellbeing), GAD-7: Generalized Anxiety Disorder Scale (Score 0-4=Minimal Anxiety, 5-9=Mild Anxiety, 10-14=Moderate Anxiety, >15=Severe Anxiety), PSS-10: Perceived Stress Scale (Score 0-13 =low stress, 14-26 = moderate stress, 27-40 =high perceived stress). High perceived stress was observed on three participants (0.3%), hence not presented in table.

The comparison of eating habits during early pandemic and the time when the response was recorded remained the same for majority of the participants (number of home cooked meals [n=186; 62%], number of meals in a day [n=182; 61%], number of snacks in a day [n=167; 56%]. Similarly, the amount of physical exercise and sleep remained the same for the majority of the participants (n=129; 43% and n=217; 72% respectively). Majority of the study participants was doing well (n=231; 77%) in terms of managing their finances and coping with their day-to-day activities (n=224; 75%).

## Discussion

The present study explored manifestations, and predictors of long COVID among the Pakistani participants. To the best of our knowledge, this is a pioneer study that has investigated the predictors and risk factors associated with the development of long COVID in Pakistan. In addition, this study has also analysed the psychosocial impact of COVID-19. Furthermore, this study has assessed the persistence of symptoms after 3 months (12 weeks) of acute COVID-19 illness. Most of the previous studies investigating the long COVID were carried out in the initial phase after recovery from acute infection (Qamar et al., 2022),(Iqbal et al., 2021).

35% of the present study participants who suffered from COVID-19 reported to have experienced symptoms beyond 12 weeks (long/post COVID disease). Qamar *et al*. conducted an online survey to assess the persistence of symptoms among COVID 19 survivors in Pakistan and reported that 83.7% of the respondents had residual symptoms after one month of the COVID 19 infection (Qamar et al., 2022). This higher rate of prevalence is most likely due to the assessment of symptoms in the early period after the acute infection. Hossain *et al*. reported that the prevalence of long COVID symptoms at 12 weeks among a cohort of Bangladeshi population was found to be 16.1% (Hossain et al., 2021). Kayaaslan *et al*. found out that 47.5% of the participants had persistent symptoms beyond 12 weeks (Kayaaslan et al., 2021). A few other studies have also reported higher rates (87.5% and 96%) of prevalence of post COVID syndrome (Leth et al., 2021),(Chopra et al., 2021). This could possibly be due to the studies being conducted in hospitalized patients and likelihood of inclusion of relatively severely ill patients in these studies. The differences in the results of various studies reporting the prevalence of long COVID can be attributed to the differences in the characteristics of the study participants (outpatient/hospitalized, varying severity of disease), duration after the acute phase of infection and data collection methods (online questionnaires, mobile apps, one to one interview, examination of outpatient and inpatient records etc.)

Muscle problems and fatigue were the most frequent (14.7%) symptoms encountered by the participants of this study beyond 12 weeks of acute COVID illness, followed by breathing problems (12.6%). Qamar *et al*. found body aches to be the commonest residual symptom followed by low mood, among a cohort of Pakistani participants (Qamar et al., 2022). Fatigue was reported to be the most prevalent persistent symptom reported in Pakistan, India and other studies conducted around different regions around the world (Iqbal et al., 2021) (Budhiraja et al., 2021) (Kayaaslan et al., 2021),(Leth et al., 2021),(Chopra et al., 2021) (Davis et al., 2021).

22.2% of the long haulers included in this study developed a new condition that might have association with COVID as diagnosed by their physician. The most prevalent among these newly diagnosed conditions was post viral fatigue (38.1%). Sørensen *et al*. also reported that 7.2% of the COVID positive participants in their study were diagnosed with at least one new condition, 6-12 months post COVID; chronic fatigue syndrome being the most frequent one (4%) (Sørensen et al., 2022).

The sufferers of long COVID also reported difficulties in performing everyday life activities with the majority reporting severe difficulty or inability to concentrate while doing something, performing day-to-day work/school, and washing the whole body. Davis *et al*. also reported that 86.2% of the working respondents of their study who suffered from long COVID felt mildly or severely unable to work (Davis et al., 2021).

The results of the present study suggest that the moderate stress was found in majority of the study participants assessed based on PSS-10 scale. Similar results were observed in other studies conducted in Pakistan and Libya (Lakhdir et al., 2022) (Jahan et al., 2021). The analysis of the GAD-7 scale showed that mild to severe level of anxiety was present in the study population (43%). Previous studies conducted across the world also had similar observations where the pandemic had profound impacts on the mental health of the individuals (Table 3).

**Table 3:** Review of the psychological impacts of COVID-19 pandemic from the literature

| Reference | Country | Sample size | Severity | Time of study |
| --- | --- | --- | --- | --- |
| **Generalized Anxiety Disorder Assessment (GAD-7)** |  |  |  |  |
| Present study | Pakistan | 300 | Minimal stress= 57.0%, Moderate stress =10.3%,<br>Severe = 7%<br>Median score= 3 | September 2021-February 2022 |
| (Matsumoto et al., 2022) | Japan, Sweden | 763 (135 COVID and 628 no covid) | Moderate-severe stress = 16.7% (non-infected),<br>15.3% infected, 40% post COVID | March-June 2021 |
| (Lee et al., 2022) | South Korea | 549 | Moderate stress-severe stress= 10.6% | November 2020 |
| (Younes et al., 2022) | Lebanon | 4397 | Median score=8 | April-May 2020 |
| (Shi et al., 2021) | China | 10,492 | Mild-Severe anxiety= 34.80% | February-August 2020 |
| (McCracken et al., 2020) | Sweden | 112,020 | None=49.4, Mild=26.4, Moderate=13.6, Severe=10.6 | May-June 2020 |
| (Terán-Pérez et al., 2021) | Mexico | 1,230 | Women (Mild=34.8%, Moderate=18.3%,<br>Severe=18%)<br>Men (Mild=38.1%, Moderate=16.9%, Severe=19%) | March-May 2020 |
| (Abdalla et al., 2021) | USA | 1,450 | Women (GAD≥10= 14%), Male (GAD≥10= 7.6%) | March-April 2020 |
| (Passos et al., 2020) | Brazil and Portugal | 550 | Mild=43.1%, Moderate=17.6%, severe=10.5% | May-July 2020 |
| **Perceived stress scale (PSS-10)** |  |  |  |  |
| Present study | Pakistan | 300 | Low stress=15.7%, Moderate stress=83.3%, | September 2021-February 2022 |
| (Lee et al., 2022) | South Korea | 549 | High stress=5.1% | November 2020 |
| (Jahan et al., 2021) | Libya | 683 | Male (low stress=41.5%, moderate= 48.9%, High=9.5%) Female (low stress=22.1%, moderate= 55.4%, High=22.6%) | NA |
| (Lakhdar et al., 2022) | Pakistan | 1654 | Low stress=16.99%, Moderate stress=68.86%, High stress=14.15% | April-August 2020 |

NA: not available. The search strategy for literature review is given in Supplementary materials

This study had several limitations. Data for clinical parameters of the study participants was not available, therefore the correlation of long COVID with clinical features could not be identified. Moreover, since the data was collected directly from the patients, the symptoms are not clinically correlated; rather the study is based on self-reported symptoms and the chances of recall bias due retrospective nature of the study are higher. Since the sample size for the long COVID cohort was small, these results do not represent the prevalence and disease burden of long COVID in Pakistan.

## Conclusion

Our results suggest that the COVID-19 illness can pose long term impacts on the health of the individuals. Fatigue and muscle aches were the most common long-term symptoms of COVID 19. Moreover, the pandemic had profound impacts on the mental health of the participants. However, extended studies to assess the prevalence and underlying factors for long COVID need to be conducted to devise an effective management plan for these individuals.

## Geolocation information

Pakistan, Southeast Asia

## Data Availability

The data for this study is not publicly available.

## Acknowledgement

We are grateful to the staff of Shifa International Hospital, Islamabad and Shifa Tameer-e-Millat University, Islamabad, Pakistan for providing facilitation in the conduction of the interviews.

## Funding details

No funding was acquired for this study.

## Disclosure statement

The authors report there are no competing interests to declare.

## Data availability statement

The data for this study is not publicly available.

## Financial support

This research did not receive any specific grant from funding agencies in the public, commercial, or not-for-profit sectors.

## Conflict of interests

None to declare.

## Data availability statement

Data sharing is not applicable to this article.

---

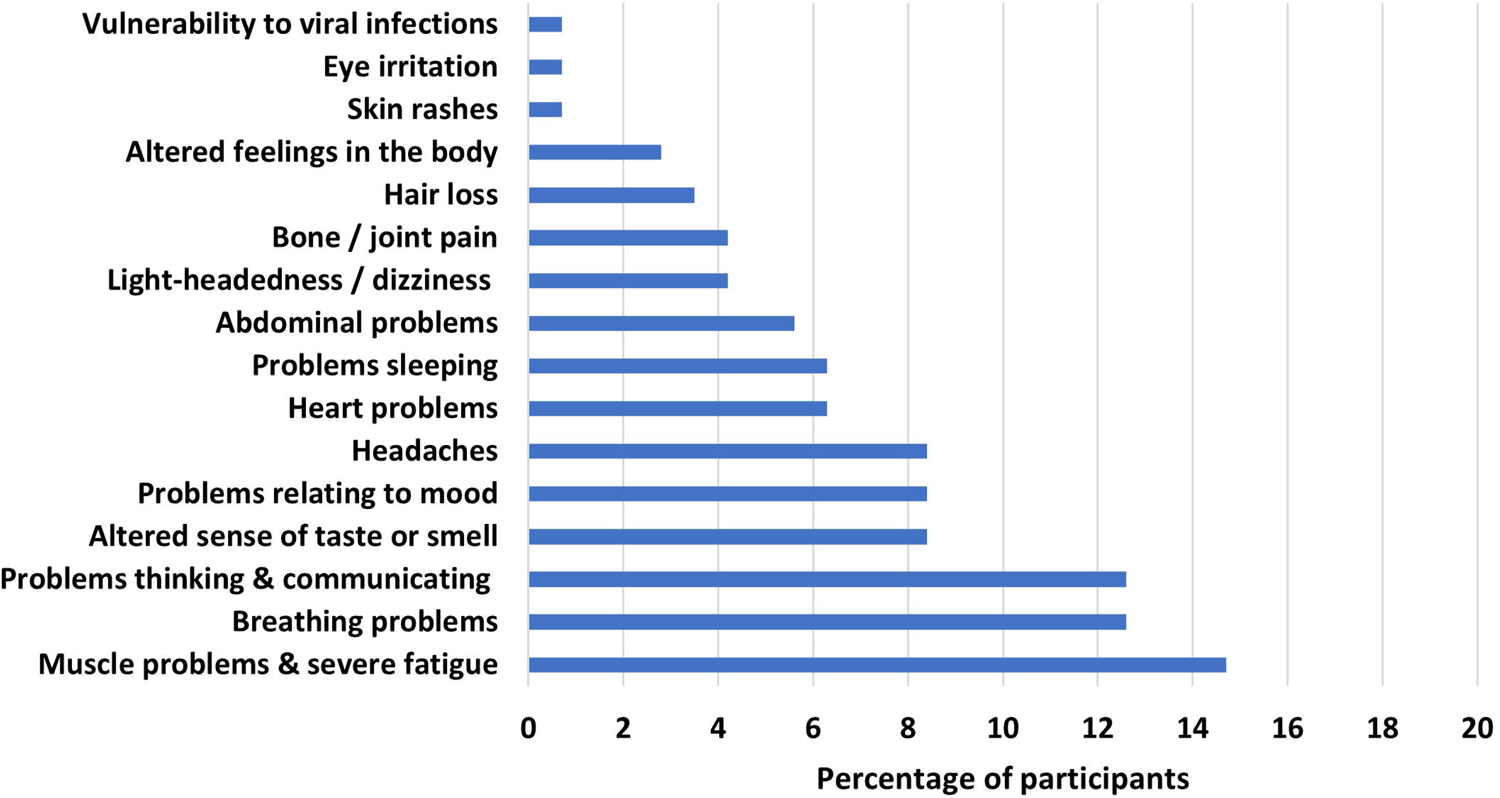

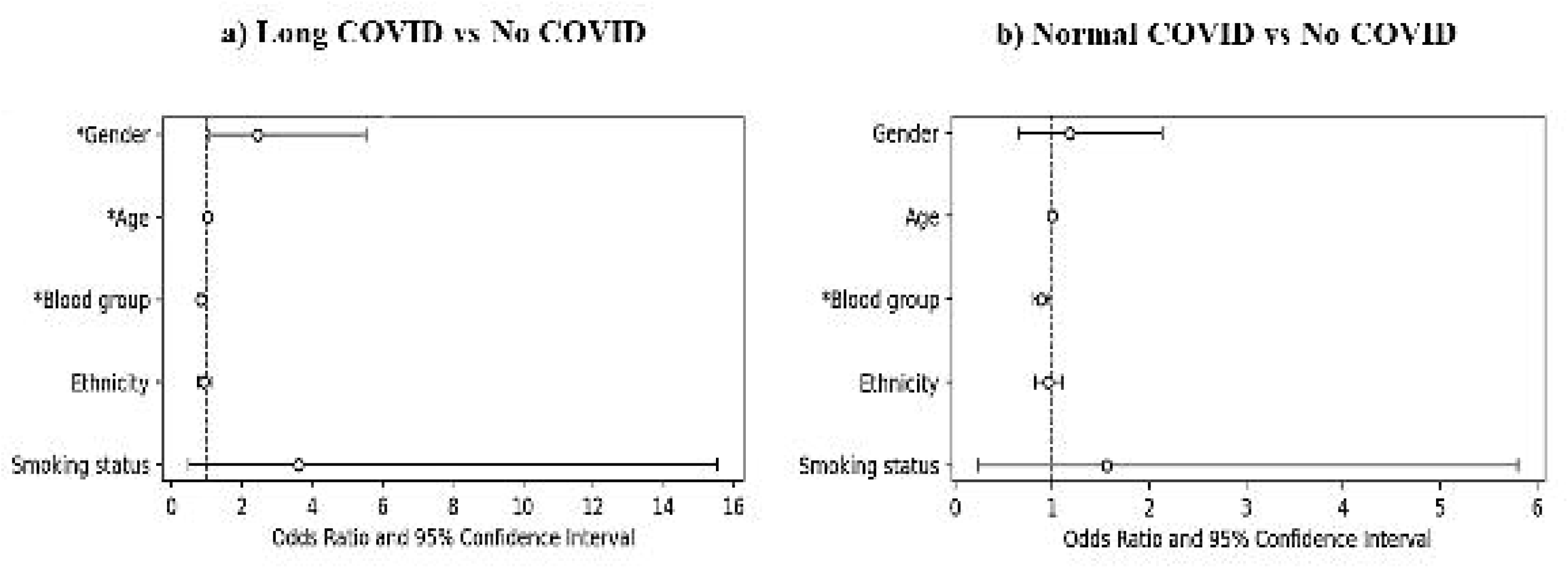

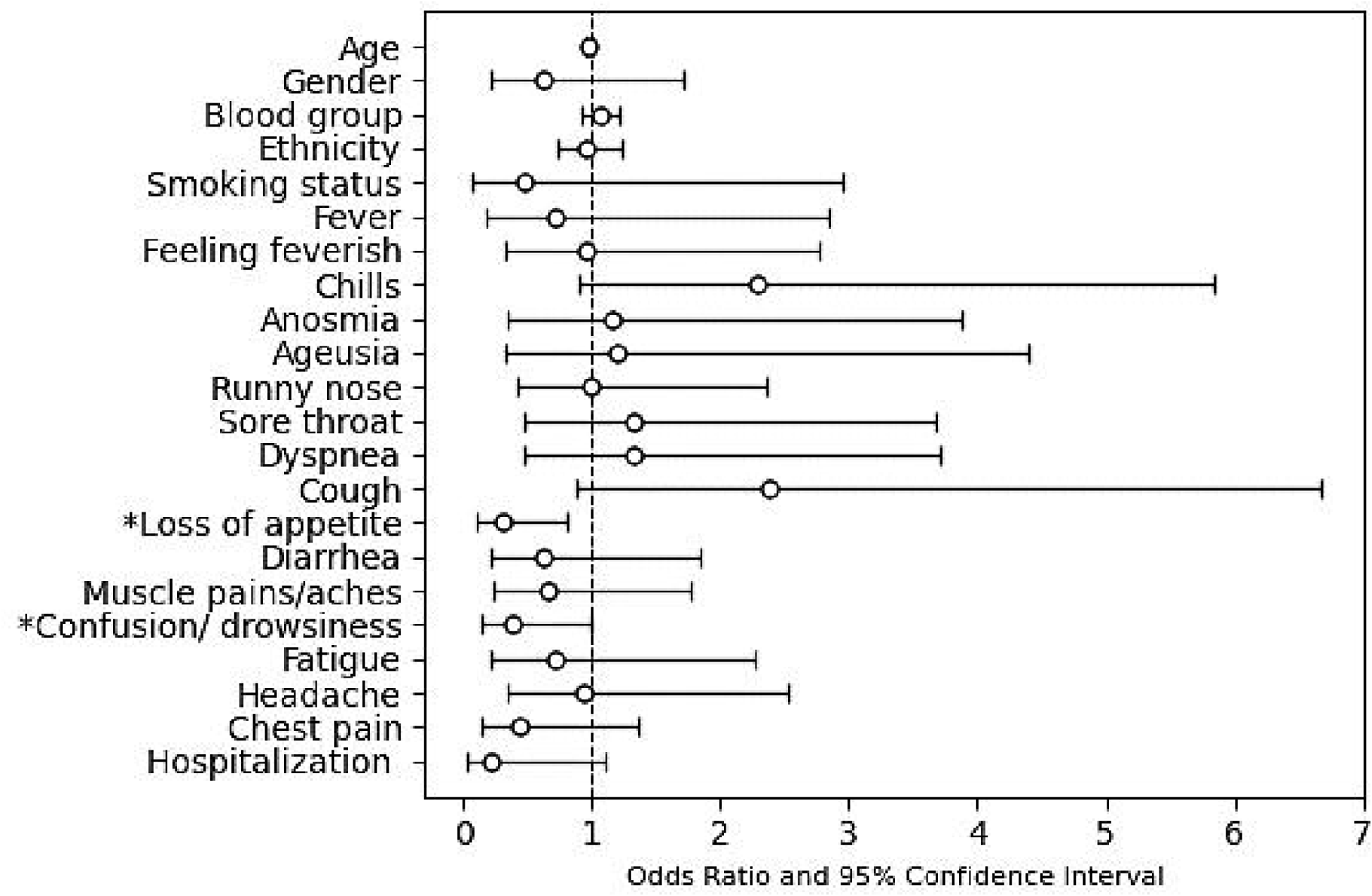

## Notes

### Competing Interest Statement

The authors have declared no competing interest.

### Author Declarations

The study was approved by Institutional Review Board & Ethics Committee (IRB&EC), Shifa Tameer-e-Millat University Islamabad (IRB # 274-21)

## References

Abdalla, S. M., Ettman, C. K., Cohen, G. H., & Galea, S. (2021). Mental health consequences of COVID-19: a nationally representative cross-sectional study of pandemic-related stressors and anxiety disorders in the USA. BMJ Open, 11(8). https://doi.org/10.1136/BMJOPEN-2020-044125

Al-Hakeim, H. K., Al-Rubaye, H. T., Al-Hadrawi, D. S., Almulla, A. F., & Maes, M. (2022). Long-COVID post-viral chronic fatigue and affective symptoms are associated with oxidative damage, lowered antioxidant defenses and inflammation: a proof of concept and mechanism study. Molecular Psychiatry 2022, 1–15. https://doi.org/10.1038/s41380-022-01836-9

Angold, A., Costello, E. J., Messer, S. C., & Pickles, A. (1995). Development of a short questionnaire for use in epidemiological studies of depression in children and adolescents. International Journal of Methods in Psychiatric Research, 5, 237–249.

Aydillo, T., Gonzalez-Reiche, A. S., Aslam, S., Guchte, A. van de, Khan, Z., Obla, A., Dutta, J., Bakel, H. van, Aberg, J., García-Sastre, A., Shah, G., Hohl, T., Papanicolaou, G., Perales, M.-A., Sepkowitz, K., Babady, N. E., & Kamboj, M. (2020). Shedding of Viable SARS-CoV-2 after Immunosuppressive Therapy for Cancer. The New England Journal of Medicine, 383(26), 2586– 2588. https://doi.org/10.1056/NEJMC2031670

Budhiraja, S., Aggarwal, M., Wig, R., Tyagi, A., Mishra, R., Mahajan, M., Kirtani, J., Tickoo, R., Bali, S., Dewan, A., Aggarwal, R., Saxena, P., Singh, N., Kumar, A., Chugh, I. M., Aneja, P., Dhall, S., Boobna, V., Arora, V., … Indrayan, A. (2021). Long Term Health Consequences of COVID-19 in Hospitalized Patients from North India: A follow up study of upto 12 months. MedRxiv, 2021.06.21.21258543. https://doi.org/10.1101/2021.06.21.21258543

Chopra, N., Chowdhury, M., Singh, A. K., MA, K., Kumar, A., Ranjan, P., Desai, D., & Wig, N. (2021). Clinical predictors of long COVID-19 and phenotypes of mild COVID-19 at a tertiary care centre in India. Drug Discoveries & Therapeutics, 15(3), 156–161. https://doi.org/10.5582/ddt.2021.01014

Cohen, S., Kamarck, T., & Mermelstein, R. (1983). A global measure of perceived stress. Journal of Health and Social Behavior, 24(4), 385–396. https://doi.org/10.2307/2136404

COVID-19 | Institute for Health Metrics and Evaluation. (n.d.). Retrieved 18 November 2022, from https://www.healthdata.org/covid

COVID-19 Health Advisory Platform by Ministry of National Health Services Regulations and Coordination. (n.d.). Retrieved 14 November 2022, from https://covid.gov.pk/

Davis, H. E., Assaf, G. S., McCorkell, L., Wei, H., Low, R. J., Re’em, Y., Redfield, S., Austin, J. P., & Akrami, A. (2021). Characterizing long COVID in an international cohort: 7 months of symptoms and their impact. EClinicalMedicine, 38. https://doi.org/10.1016/j.eclinm.2021.101019

GISAID - gisaid.org. (n.d.). Retrieved 18 November 2022, from https://gisaid.org/

Gupta, A., Madhavan, M. v., Sehgal, K., Nair, N., Mahajan, S., Sehrawat, T. S., Bikdeli, B., Ahluwalia, N., Ausiello, J. C., Wan, E. Y., Freedberg, D. E., Kirtane, A. J., Parikh, S. A., Maurer, M. S., Nordvig, A. S., Accili, D., Bathon, J. M., Mohan, S., Bauer, K. A., … Landry, D. W. (2020). Extrapulmonary manifestations of COVID-19. Nature Medicine, 26(7), 1017–1032. https://doi.org/10.1038/S41591-020-0968-3

Hossain, M. A., Hossain, K. M. A., Saunders, K., Uddin, Z., Walton, L. M., Raigangar, V., Sakel, M., Shafin, R., Hossain, M. S., Kabir, M. F., Faruqui, R., Rana, M. S., Ahmed, M. S., Chakrovorty, S. K., Hossain, M. A., & Jahid, I. K. (2021). Prevalence of Long COVID symptoms in Bangladesh: a prospective Inception Cohort Study of COVID-19 survivors. BMJ Global Health, 6(12), e006838. https://doi.org/10.1136/BMJGH-2021-006838

Iqbal, A., Iqbal, K., Ali, S. A., Azim, D., Farid, E., Baig, M. D., Arif, T. bin, & Raza, M. (2021). The COVID-19 Sequelae: A Cross-Sectional Evaluation of Post-recovery Symptoms and the Need for Rehabilitation of COVID-19 Survivors. Cureus, 13(2). https://doi.org/10.7759/CUREUS.13080

Jahan, A. M., Mohamed, M., Alfagieh, M., Alnawy, N., Alsabiri, M., Algazal, R., Saaleh, R., El Swisy, S., Abbas, O., Al Delawi, W., Abdulhafith, B., Almangoush, O., Elhag, F., Elshukri, A., Abushaala, W., Shahrani, T., Tnton, A., Alkilani, H., & Dier, A. (2021). Psychosocial Impact of COVID-19 Pandemic in Libya: A Cross-Sectional Study. Frontiers in Psychology, 12, 3515. https://doi.org/10.3389/FPSYG.2021.714749/BIBTEX

Kayaaslan, B., Eser, F., Kalem, A. K., Kaya, G., Kaplan, B., Kacar, D., Hasanoglu, I., Coskun, B., & Guner, R. (2021). Post-COVID syndrome: A single-center questionnaire study on 1007 participants recovered from COVID-19. Journal of Medical Virology, 93(12), 6566–6574. https://doi.org/10.1002/jmv.27198

Lakhdir, M. P. A., Peerwani, G., Azam, S. I., Ali Nathwani, A., Iqbal, R., & Asad, N. (2022). Original research: Burden and factors associated with perceived stress amidst COVID-19: a population web-based study in Pakistan. BMJ Open, 12(6), 58234. https://doi.org/10.1136/BMJOPEN-2021-058234

Lee, H., Choi, D., & Lee, J. J. (2022). Depression, anxiety, and stress in Korean general population during the COVID-19 pandemic. Epidemiology and Health, 44. https://doi.org/10.4178/EPIH.E2022018

Leth, S., Gunst, J. D., Mathiasen, V., Hansen, K., Søgaard, O., Østergaard, L., Jensen-Fangel, S., Storgaard, M., & Agergaard, J. (2021). Persistent Symptoms in Patients Recovering From COVID-19 in Denmark. Open Forum Infectious Diseases, 8(4). https://doi.org/10.1093/OFID/OFAB042

Lopez-Leon, S., Wegman-Ostrosky, T., Perelman, C., Sepulveda, R., Rebolledo, P. A., Cuapio, A., & Villapol, S. (2021). More than 50 long-term effects of COVID-19: a systematic review and meta-analysis. Scientific Reports 2021 11:1, 11(1), 1–12. https://doi.org/10.1038/s41598-021-95565-8

Löwe, B., Decker, O., Müller, S., Brähler, E., Schellberg, D., Herzog, W., & Herzberg, P. Y. (2008). Validation and standardization of the generalized anxiety disorder screener (GAD-7) in the general population. Medical Care, 46(3), 266–274. https://doi.org/10.1097/MLR.0B013E318160D093

Matsumoto, K., Hamatani, S., Shimizu, E., Käll, A., & Andersson, G. (2022). Impact of post-COVID conditions on mental health: a cross-sectional study in Japan and Sweden. BMC Psychiatry, 22(1), 1–13. https://doi.org/10.1186/S12888-022-03874-7/TABLES/3

McCracken, L. M., Badinlou, F., Buhrman, M., & Brocki, K. C. (2020). Psychological impact of COVID-19 in the Swedish population: Depression, anxiety, and insomnia and their associations to risk and vulnerability factors. European PsychiatrylJ: The Journal of the Association of European Psychiatrists, 63(1). https://doi.org/10.1192/J.EURPSY.2020.81

Munblit, D., Nicholson, T. R., Needham, D. M., Seylanova, N., Parr, C., Chen, J., Kokorina, A., Sigfrid, L., Buonsenso, D., Bhatnagar, S., Thiruvengadam, R., Parker, A. M., Preller, J., Avdeev, S., Klok, F. A., Tong, A., Diaz, J. v., Groote, W. de, Schiess, N., … Williamson, P. R. (2022). Studying the post-COVID-19 condition: research challenges, strategies, and importance of Core Outcome Set development. BMC Medicine, 20(1), 1–13. https://doi.org/10.1186/S12916-021-02222-Y/TABLES/2

Nittas, V., Gao, M., West, E. A., Ballouz, T., Menges, D., Wulf Hanson, S., & Puhan, M. A. (2022). Long COVID Through a Public Health Lens: An Umbrella Review. Public Health Reviews, 43. https://doi.org/10.3389/PHRS.2022.1604501/FULL

Passos, L., Prazeres, F., Teixeira, A., & Martins, C. (2020). Impact on Mental Health Due to COVID-19 Pandemic: Cross-Sectional Study in Portugal and Brazil. International Journal of Environmental Research and Public Health, 17(18), 1–13. https://doi.org/10.3390/IJERPH17186794

Proal, A. D., & VanElzakker, M. B. (2021). Long COVID or Post-acute Sequelae of COVID-19 (PASC): An Overview of Biological Factors That May Contribute to Persistent Symptoms. Frontiers in Microbiology, 12, 1494. https://doi.org/10.3389/FMICB.2021.698169/XML/NLM

Qamar, M. A., Martins, R. S., Dhillon, R. A., Tharwani, A., Irfan, O., Suriya, Q. F., Rizwan, W., Khan, J. A., & Zubairi, A. bin S. (2022). Residual symptoms and the quality of life in individuals recovered from COVID-19 infection: A survey from Pakistan. Annals of Medicine and Surgery, 75. https://doi.org/10.1016/j.amsu.2022.103361

Raveendran, A. v., Jayadevan, R., & Sashidharan, S. (2021). Long COVID: An overview. Diabetes & Metabolic Syndrome: Clinical Research & Reviews, 15(3), 869–875. https://doi.org/10.1016/J.DSX.2021.04.007

Shi, L., Lu, Z. A., Que, J. Y., Huang, X. L., Lu, Q. D., Liu, L., Zheng, Y. B., Liu, W. J., Ran, M. S., Yuan, K., Yan, W., Sun, Y. K., Sun, S. W., Shi, J., Kosten, T., Bao, Y. P., & Lu, L. (2021). Long-term impact of covid-19 on mental health among the general public: A nationwide longitudinal study in China. International Journal of Environmental Research and Public Health, 18(16), 8790. https://doi.org/10.3390/IJERPH18168790/S1

Smith, D., Bowring, C., Wells, N., Crawford, M., Timpson, N. J., & Northstone, K. (2021). The Avon Longitudinal Study of Parents and Children - A resource for COVID-19 research: Questionnaire data capture November 2020L? March 2021 [version 2; peer review: 2 approved]. Wellcome Open Research, 6(155). https://doi.org/10.12688/wellcomeopenres.16950.2

Sørensen, A. I. V., Spiliopoulos, L., Bager, P., Nielsen, N. M., Hansen, J. V., Koch, A., Meder, I. K., Ethelberg, S., & Hviid, A. (2022). A nationwide questionnaire study of post-acute symptoms and health problems after SARS-CoV-2 infection in Denmark. Nature Communications 2022 13:1, 13(1), 1–8. https://doi.org/10.1038/s41467-022-31897-x

Tennant, R., Hiller, L., Fishwick, R., Platt, S., Joseph, S., Weich, S., Parkinson, J., Secker, J., & Stewart-Brown, S. (2007). The Warwick-Dinburgh mental well-being scale (WEMWBS): Development and UK validation. Health and Quality of Life Outcomes, 5(1), 1–13. https://doi.org/10.1186/1477-7525-5-63/TABLES/3

Terán-Pérez, G., Portillo-Vásquez, A., Arana-Lechuga, Y., Sánchez-Escandón, O., Mercadillo-Caballero, R., González-Robles, R. O., & Velázquez-Moctezuma, J. (2021). Sleep and Mental Health Disturbances Due to Social Isolation during the COVID-19 Pandemic in Mexico. International Journal of Environmental Research and Public Health, 18(6), 1–11. https://doi.org/10.3390/IJERPH18062804

WHO Coronavirus (COVID-19) Dashboard | WHO Coronavirus (COVID-19) Dashboard With Vaccination Data. (2022). Retrieved 14 November 2022, from https://covid19.who.int/

Younes, S., Safwan, J., Rahal, M., Hammoudi, D., Akiki, Z., & Akel, M. (2022). Effect of COVID-19 on mental health among the young population in Lebanon. L’Encephale, 48(4), 371–382. https://doi.org/10.1016/J.ENCEP.2021.06.007

ZOE Health Study. (2022). Retrieved 11 November 2022, from https://health-study.joinzoe.com/

