## Supplementary Material for "Analysing the psychosocial and health impacts of Long COVID in Pakistan: A cross sectional study"

**Section 1. Tables**

**Supplementary Table 1: Characteristics of COVID-19 positive patients**

| Characteristics | Overall (n=155) | Normal COVID (n=101) | Long COVID (n=54) |
|---|---|---|---|
| **Medical help for COVID symptoms** | | | |
| Visited doctor/GP/Nurse | 113 (73) | 66 (65) | 47 (87.0) |
| Visited pharmacist | 7 (4.5) | 6 (6) | 1 (1.9) |
| No medical help sought | 25 (23) | 29 (29) | 6 (11.1) |
| **Hospitalized due to COVID symptoms** | | | |
| Yes | 13 (8.4) | 4 (4) | 9 (16.7) |
| No | 142 (91.6) | 97 (96) | 45 (83.3) |
| **Medications for COVID symptoms** | | | |
| Antipyretics | 102 (66) | 61 (60.3) | 41 (76) |
| Antibiotics | 60 (39) | 31 (31) | 31 (57) |
| Steroids | 14 (9) | 5 (5) | 9 (17) |
| Multivitamins | 98 (63) | 53 (52) | 45 (83) |
| Remdesivir | 5 (3.2) | 2 (2) | 3 (5) |
| Anticoagulants | 14 (9) | 6 (6) | 8 (15) |
| Home remedies | 64 (41.3) | 34 (34) | 30 (56) |
| No treatment/Unknown | 27 (17.4) | 19 (19) | 8 (15) |
| Anti-Allergy | 5 (3.2) | 4 (4) | 1 (2) |
| **Bedridden due to overall symptoms** | | | |
| Never | 15 (9.7) | 11 (10.8) | 4 (7.4) |
| For a week | 86 (28.6) | 65 (64.3) | 20 (37.1) |
| 1 to 3 weeks | 46 (15.3) | 25 (24.7) | 22 (40.8) |
| 4 to 12 weeks | 5 (3.2) | 0 (0.0) | 5 (9.3) |
| More than 12 weeks | 3 (1.9) | 0 (0.0) | 3 (5.6) |

**Supplementary Table 2: List of new conditions diagnosed among the Long COVID patients**

| Characteristics | Percentage |
|---|---|
| **New condition linked to COVID-19 indicated by a physician** | |
| Yes | 22.2 |
| No | 77.8 |
| **New condition diagnosed (%)** | |
| Post-viral fatigue | 38.1 |
| Post-COVID syndrome | 14.3 |
| A blood clot in the leg, lung, heart or brain | 4.8 |
| A heart condition | 4.8 |
| A lung condition | 9.5 |

| | |
|---|---|
| A condition affecting the nervous system outside the brain | 4.8 |
| A condition affecting the kidneys | 4.8 |
| Thyroid disease | 4.8 |
| Chronic utricaria | 4.8 |
| Eye floaters | 4.8 |
| Eosinophilic Colitis | 4.8 |

**Supplmentary table 3: Comparison of psychological impacts of COVID-19 pandemic among different groups**

| | N | Median | IQR | p-value |
|---|---|---|---|---|
| **Short Mood and feeling questionnaire (sMFQ)** | | | | |
| Overall | 300 | 2 | 9.00-0.00 | |
| No COVID | 145 | 2 | 8.00-0.00 | 0.00 |
| Normal COVID | 101 | 2 | 8.00-0.00 | |
| Long COVID | 54 | 7 | 9.50-1.75 | |
| **Warwick-Edinburgh Mental Well-being Scale (WEMWBS)** | | | | |
| Overall | 300 | 51 | 55.00-43.00 | |
| No COVID | 145 | 51 | 55.00-43.50 | 0.13 |
| Normal COVID | 101 | 50 | 55.00-42.00 | |

| | | | | |
|---|---|---|---|---|
| Long COVID | 54 | 47 | 54.00-42.00 | |
| **Generalized Anxiety Disorder Assessment (GAD-7)** | | | | |
| Overall | 300 | 3 | 7.00-1.00 | |
| No COVID | 145 | 2 | 6.00-1.00 | 0.06 |
| Normal COVID | 101 | 3 | 7.00-1.00 | |
| Long COVID | 54 | 5 | 9.70-0.00 | |
| **Perceived Stress Scale (PSS)** | | | | |
| Overall | 300 | 18 | 22.00-16.00 | |
| No COVID | 145 | 18 | 21.00-16.00 | 0.25 |
| Normal COVID | 101 | 16 | 22.00-13.00 | |
| Long COVID | 54 | 19.5 | 23.00-15.75 | |

### Section 2. Figures

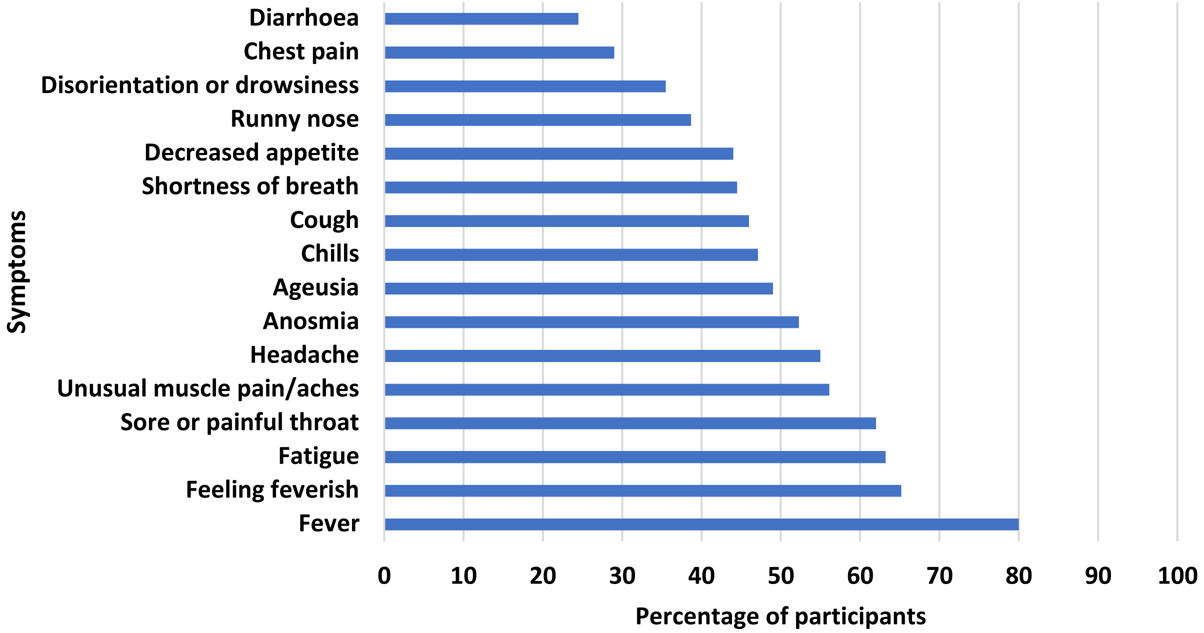


**Supplementary Figure 1: Symptoms reported during the acute phase of COVID-19 illness by the study participants (n=155)**

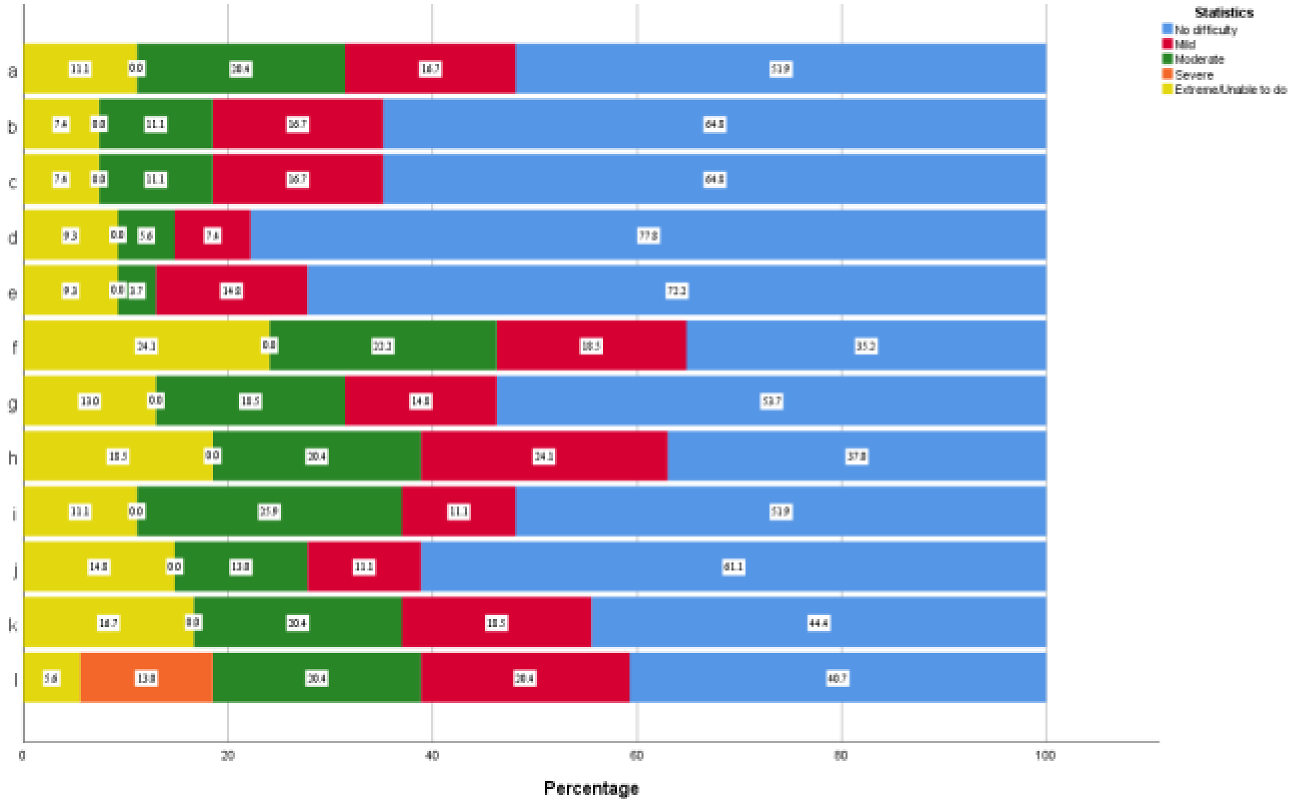


**Supplementary Figure 2: Difficulty levels in daily life activities 12 weeks after COVID-19 illness;** a) Your day-to-day work/school b) Maintaining a friendship c) Dealing with people you do not know d)

Getting dressed e) Washing your whole body f) Walking a long distance such as 1 kilometre or half a mile g) Concentrating on doing something for ten minutes h) Being emotionally affected by your health problems i) Joining in community activities (e.g. festivities, religious, other) j) Learning a new task, e.g. learning how to get to a new place k) Taking care of your household responsibilities l) Standing for long periods such as 30 minutes

**Section 3. Questionnaire**

**This questionnaire consisted of 5 sections:**

A – Personal information and medical history of the respondants and their general health before the pandemic
B – COVID-19 illness, illness history (hospitalization, duration) and treatment
C – COVID-19 testing
D – Vaccination for COVID-19
E – Social and pscyhcological imapcts of COVID-19 pandemic

### A: Personal Information and medical history

**Q1:**

a) Name of the particpant
b) Age
c) Gender
d) Area of residence
e) Ethnicity
f) Blood group

**Q2. In general, in the <u>3 months before the COVID-19 outbreak in March 2020</u>, would you say your health was...?**

- ◻ Excellent
- ◻ Very good
- ◻ Good
- ◻ Fair
- ◻ Poor
- ◻ Don’t know
- ◻ Prefer not to say

**Q3. Were you contacted by letter or text message to say you are at severe risk from COVID-19 due to an underlying health condition and should be shielding?**

- ◻ Yes
- ◻ No

**3A. Please specify :**

**Q4. Do you smoke?**

- ◻ Yes
- ◻ No

### B: COVID-19 status

**Q5. Do you think that you currently have or have ever had COVID-19?**
- ◻ Yes, confirmed by a positive test
- ◻ Yes, based on medical advice
- ◻ Yes, based on strong personal suspicion
- ◻ Unsure
- ◻ No → **Go to Section C**
- ◻ Don’t know → **Go to Section C**
- ◻ Prefer not to say → **Go to Section C**

**Q6. When do you think you first got (or might have got) COVID-19? If you do not remember exactly, please put your best estimate.**

DD/MM/YYYY __ __ /__ __ /__ __ __ __
- ◻ Don’t know
- ◻ Prefer not to answer

**Q7. In this question we would like to know if you have had ANY of the following symptoms from ANY illness you have had.**
- ◻ Fever
- ◻ Feeling feverish
- ◻ Chills (feeling too cold)
- ◻ Loss or change in sense of smell
- ◻ Loss or change in sense of taste
- ◻ Runny nose
- ◻ Sore or painful throat
- ◻ Shortness of breath or trouble breathing affecting normal activities
- ◻ New persistent cough
- ◻ Decrease in appetite
- ◻ Diarrhoea
- ◻ Unusual muscle pains or aches
- ◻ Confusion, disorientation or drowsiness
- ◻ Unusual fatigue/ feeling unusually tired
- ◻ Headache
- ◻ Chest pain
- ◻ Other:

**Q8. In the first 4 weeks of illness, did you look for any medical help for any symptoms you think may have been caused by COVID-19?**
- ◻ Yes – discussed symptoms with doctor/GP/practice nurse
- ◻ Yes – visited pharmacist
- ◻ Yes – visited A&E or walk in centre
- ◻ No

- ◻ Don’t know
- ◻ Prefer not to say

**Q9. Did you look for any medical help for any symptoms you had more than 4 weeks after your symptoms began, that you think may have been caused by COVID-19?**
*Please select all that apply.*

- ◻ Yes – discussed symptoms with doctor/GP/practice nurse
- ◻ Yes – visited pharmacist
- ◻ Yes – visited A&E or walk in centre
- ◻ Yes – outpatient consultation for long Covid
- ◻ No
- ◻ Don’t know
- ◻ Prefer not to say

**Q10. Have you ever had to stay in hospital because of COVID-19 symptoms?**

- ◻ Yes
- ◻ No
- ◻ Don’t know
- ◻ Prefer not to say

**Q11. Do you think you have caught COVID-19 more than once?**

- ◻ Yes, confirmed by a second positive test
- ◻ Yes, based on medical advice
- ◻ Yes, based on strong personal suspicion
- ◻ Unsure
- ◻ No
- ◻ Don’t know
- ◻ Prefer not to say

**Q12. When did you catch COVID-19 the second time? If you do not remember exactly, please put your best estimate.**

DD/MM/YYYY __ __ /__ __ /__ __ __ __

**Q13. When did you catch COVID-19 the second time? If you do not remember exactly, please put your best estimate.**
DD/MM/YYYY __ __ /__ __ /__ __ __ __

**Q14. Thinking of your last, or only episode of COVID-19, have you now recovered to normal?**

- ◻ Yes, I am back to normal
- ◻ No, I still have some or all of my symptoms

**Q15. How long have you had / did you have COVID-19 symptoms overall? Please include time spent with mild symptoms and the time in between symptoms if these have been coming and going.**
If you have caught COVID-19 more than once, please answer about the longest episode of illness you experienced.

- ◻ Less than 2 weeks
- ◻ 2-3 weeks
- ◻ 4-12 weeks
- ◻ More than 12 weeks

**Q16. For how long were you or have you been unable to function as normal due to COVID-19 symptoms?**

- ◻ I was always able to function as normal
- ◻ 1-3 days
- ◻ 4-6 days
- ◻ 7-13 days
- ◻ 2-3 weeks
- ◻ 4-12 weeks
- ◻ 12+ weeks

**Q17. How many days were you or have you been so unwell that you stayed in bed or on the sofa?**

- ◻ None
- ◻ 1-3 days
- ◻ 4-6 days
- ◻ 7-13 days
- ◻ 2-3 weeks
- ◻ 4-12 weeks
- ◻ 12+ weeks

**Q18. Which of the following remedies did you use either on doctor's prescription or on your own?**

- ◻ Surbex - Z
- ◻ Vitamin D (Injection/ Gel capsule)
- ◻ Oral Azithromycin
- ◻ IV Azithromycin
- ◻ Oral Levofloxacin
- ◻ IV Levofloxacin
- ◻ IV Meropenem
- ◻ IV Piperacillin
- ◻ IV Fluconazole
- ◻ IV Vancomycin
- ◻ IV Remdesivir
- ◻ Oral Prednisolone
- ◻ IV Prednisolone
- ◻ Oral Dexamethasone
- ◻ IV Dexamethasone
- ◻ Oral Panadol
- ◻ IV Panadol
- ◻ Enflor sachet
- ◻ Oral Anticoagulants: (Loprin, enoxaparin, apixaban)
- ◻ IV anticoagulant (Clexane)
- ◻ Home remedies
- ◻ Any other

If you have used any other remedy please specify

______________________________________________

**Q19. Have you been told by a doctor that you may have a new condition, illness, or disability because of infection with COVID-19?**

- ◻ Yes
- ◻ No

**Q20. Please indicate what new condition, illness or disability your doctor has linked to COVID-19.**
*Please tick all that apply*

- ◻ Post-viral fatigue
- ◻ Post-COVID syndrome
- ◻ A blood clot in the leg, lung, heart or brain
- ◻ A heart condition
- ◻ A lung condition
- ◻ A condition affecting the mind or brain
- ◻ A condition affecting the nervous system outside the brain
- ◻ A condition affecting the kidneys
- ◻ Thyroid disease
- ◻ Other – please specify _____________

**Q21. Did you have any of the following problems 12 weeks (or more) after first catching COVID-19? Please only consider symptoms that are not explained by another reason.** *Tick all that apply.*

- ◻ I was back to my usual self  **Go to Section C**
- ◻ Breathing problems, e.g. breathlessness, pain on breathing, cough
- ◻ Altered sense of taste or smell
- ◻ Problems thinking and communicating e.g. brain-fog, memory problems, difficulty concentrating, decreased alertness, confusion, difficulty speaking
- ◻ Heart problems, e.g. chest pain, palpitations
- ◻ Light-headedness / dizziness on standing
- ◻ Abdominal problems e.g. tummy pain, diarrhoea, appetite loss
- ◻ Muscle problems, e.g. muscle aches, weakness, severe fatigue
- ◻ Altered feelings in your body, e.g. unusual tingling, pain
- ◻ Problems relating to mood, e.g. anxiety, feeling 'down', or irritable
- ◻ Problems sleeping, e.g. poor sleep or excessive sleep
- ◻ Skin rashes
- ◻ Bone / joint pain
- ◻ Headaches
- ◻ Other:

**Q22. How much difficulty did you have with the following activities 12 weeks (3 months) after your COVID-19 illness began?**

| | **No difficulty** | **Mild** | **Moderate** | **Severe** | **Extreme/Unable to do** |
|---|---|---|---|---|---|

| | | | | | |
|---|---|---|---|---|---|
| Standing for long periods such as 30 minutes? | ◻ | ◻ | ◻ | ◻ | ◻ |
| Taking care of your household responsibilities? | ◻ | ◻ | ◻ | ◻ | ◻ |
| Learning a new task, e.g. learning how to get to a new place? | ◻ | ◻ | ◻ | ◻ | ◻ |
| Joining in community activities (e.g. festivities, religious, other)? | ◻ | ◻ | ◻ | ◻ | ◻ |
| Being emotionally affected by your health problems? | ◻ | ◻ | ◻ | ◻ | ◻ |
| Concentrating on doing something for ten minutes? | ◻ | ◻ | ◻ | ◻ | ◻ |
| Walking a long distance such as 1 kilometre or half a mile? | ◻ | ◻ | ◻ | ◻ | ◻ |
| Washing your whole body? | ◻ | ◻ | ◻ | ◻ | ◻ |
| Getting dressed? | ◻ | ◻ | ◻ | ◻ | ◻ |
| Dealing with people you do not know? | ◻ | ◻ | ◻ | ◻ | ◻ |
| Maintaining a friendship? | ◻ | ◻ | ◻ | ◻ | ◻ |
| Your day-to-day work/school? | ◻ | ◻ | ◻ | ◻ | ◻ |

**Q23. Thinking of how you felt 12 weeks after your COVID-19 illness began, what did you need help with because of COVID-19?** *Please select all that apply.*

- ◻ Getting essential shopping, e.g. food or medication
- ◻ Preparing food and/or drink
- ◻ Washing and dressing

- ◻ Housework, e.g. laundry, cleaning or hoovering
- ◻ Managing household responsibilities, e.g. finances or paying bills
- ◻ Day to day work / study
- ◻ Childcare or other caring responsibilities
- ◻ Letting other people know about my illness (e.g. employer, university, family)
- ◻ Getting about (travel), e.g. driving
- ◻ I have not needed any additional support

**Q24. What help or support have you found helpful, 12 weeks after your COVID-19 illness began?**
*Please select all that apply.*

- ◻ Self-organized group or network of people with the same condition e.g. on social media
- ◻ Local volunteer network
- ◻ Support from people you live with
- ◻ Support from neighbours
- ◻ Support from a religious group
- ◻ Support from a charity
- ◻ Support from family
- ◻ Support from friends
- ◻ Support from your local council
- ◻ Support from your GP
- ◻ I'm not sure what would be most helpful

**Q25. What help do you think would be most useful for people who continue to have symptoms 12 weeks after their COVID-19 illness began?** *Please select the top three most useful.*

- ◻ Network of people with the same condition
- ◻ Reliable, easily accessible information in one place
- ◻ Access to financial support
- ◻ Access to supermarket / food deliveries
- ◻ Access to therapy – e.g. occupational or physical therapy
- ◻ Access to psychological support
- ◻ Access to a doctor and if necessary, specialist care
- ◻ I'm not sure what would be most useful

### C: Testing for Coronavirus

**Q26. Have you ever had a swab test to see if you have COVID-19 (of your nose and/or throat, or saliva)?** *Please select all that apply.*

- ◻ Yes, because of my job / studying (e.g. routine swab tests)
- ◻ Yes, because I had symptoms
- ◻ Yes, because I had been in contact with someone who had COVID-19
- ◻ Yes, because I have taken part in a research study
- ◻ Yes, because of travel
- ◻ Yes, because I needed a medical procedure (not related to COVID-19)
- ◻ Yes, because my local area was involved in routine swabbing
- ◻ Yes, other
- ◻ No

**Q27. Can you provide the dates of your swab / saliva tests and results?** *If you can't remember exactly, please give your best estimate*

DD/MM/YYYY __ __ /__ __ /__ __ __ __ Positive □ Negative □ Unknown □ Prefer not to say □
DD/MM/YYYY __ __ /__ __ /__ __ __ __ Positive □ Negative □ Unknown □ Prefer not to say □
DD/MM/YYYY __ __ /__ __ /__ __ __ __ Positive □ Negative □ Unknown □ Prefer not to say □
DD/MM/YYYY __ __ /__ __ /__ __ __ __ Positive □ Negative □ Unknown □ Prefer not to say □
DD/MM/YYYY __ __ /__ __ /__ __ __ __ Positive □ Negative □ Unknown □ Prefer not to say □
DD/MM/YYYY __ __ /__ __ /__ __ __ __ Positive □ Negative □ Unknown □ Prefer not to say □

**Q28. Can you provide the dates of your tests and results where they were positive (if any)?** Please include all positive test results you have received, whether due to routine testing or for any other reason. Do not include negative or inconclusive results. *If you can't remember exactly, please give your best estimate.*

DD/MM/YYYY __ __ /__ __ /__ __ __ __ Positive □
DD/MM/YYYY __ __ /__ __ /__ __ __ __ Positive □
DD/MM/YYYY __ __ /__ __ /__ __ __ __ Positive □
DD/MM/YYYY __ __ /__ __ /__ __ __ __ Positive □
DD/MM/YYYY __ __ /__ __ /__ __ __ __ Positive □

**Q29. When did you start routine testing for your work / study?** *If you can't remember exactly, please give your best estimate.*

DD/MM/YYYY __ __ /__ __ /__ __ __ __ Don't know □

**Q30. Have you ever had a blood or finger-prick test to see if you had past infection with COVID-19 (sometimes called antibody or serology tests)?** Please select all that apply.

- □ Yes, because I previously had symptoms
- □ Yes, because I took part in a research study
- □ Yes, because of my job / studying (e.g. routine antibody tests)
- □ Yes, other
- □ No
- □ Don't know

**Q31. Can you provide the dates of your blood or finger-prick tests and results?** *If you can't remember exactly, please give your best estimate.*

DD/MM/YYYY __ __ /__ __ /__ __ __ __ Positive □ Negative □ Unknown □
DD/MM/YYYY __ __ /__ __ /__ __ __ __ Positive □ Negative □ Unknown □
DD/MM/YYYY __ __ /__ __ /__ __ __ __ Positive □ Negative □ Unknown □
DD/MM/YYYY __ __ /__ __ /__ __ __ __ Positive □ Negative □ Unknown □
DD/MM/YYYY __ __ /__ __ /__ __ __ __ Positive □ Negative □ Unknown □
DD/MM/YYYY __ __ /__ __ /__ __ __ __ Positive □ Negative □ Unknown □

### D: Vaccination

**Q32. Have you been invited to have a COVID-19 vaccine?**

- ◻ Yes
- ◻ No → **Go to Section E**
- ◻ Don’t know → **Go to Section E**
- ◻ Prefer not to say → **Go to Section E**

**Q33. Have you had at least one COVID-19 vaccine injection?**

- ◻ Yes
- ◻ No – but I intend to → **Go to Section E**
- ◻ No – and I do not intend to → **Go to Section E**
- ◻ Don’t know → **Go to Section E**
- ◻ Prefer not to say → **Go to Section E**

**Q34. What is the name of the vaccine you received?**

- ◻ Oxford AstraZeneca
- ◻ Pfizer BioNTech
- ◻ Sputnik V
- ◻ Sinopharm
- ◻ SinoVac
- ◻ Cansino
- ◻ PakVac
- ◻ Other – please specify________
- ◻ Don’t know

**Q35. When was your first COVID-19 vaccine injection**? *If you can’t remember exactly, please put your best estimate.*

DD/MM/YYYY __ __ /__ __ /__ __ __ __

- ◻ Don’t know

**Q36. Have you had your second COVID-19 vaccine injection yet?**

- ◻ No
- ◻ Yes

**Q37. What is the name of the second dose vaccine you received?**

- ◻ Oxford AstraZeneca
- ◻ Pfizer BioNTech
- ◻ Sputnik V
- ◻ Sinopharm
- ◻ Sinovac
- ◻ PakVac
- ◻ Cansino
- ◻ Other – please specify__________
- ◻ Don’t know

**Q38. When was your second COVID-19 vaccine injection?** *If you can’t remember exactly, please put your best estimate.*

DD/MM/YYYY __ __ /__ __ /__ __ __ __

- ◻ Don’t know

**Q39. Have you had booster dose of vaccine?**

- ◻ Yes
- ◻ No

**Q40. What is the name of booster dose you received?**

- ◻ Sinopharm
- ◻ Sinovac
- ◻ Pfizer
- ◻ Moderna
- ◻ Other-please specify

**Q41. When was your booster COVID-19 vaccine injection?** *If you can’t remember exactly, please put your best estimate.*

DD/MM/YYYY __ __ /__ __ /__ __ __ __

**Q42. Did you have ongoing symptoms from COVID-19 in the week before you were given your first COVID-19 vaccine injection?**

- ◻ Yes
- ◻ No → **Go to Section E**
- ◻ Don’t know → **Go to Section E**

**Q43. Please tell us whether your symptoms from your COVID-19 illness changed 2 weeks or later after having your first COVID-19 vaccine injection.**

- ◻ Yes – They all got better
- ◻ Yes – Some of them got better
- ◻ No change
- ◻ Yes – Some of them got worse
- ◻ Yes – They all got worse
- ◻ Some improved and others got worse
- ◻ It has not yet been 2 weeks since my first COVID-19 vaccine injection

### E: Questions about coronavirus/COVID-19 and its effect on your life

**Q44.Thinking about life now, compared to the early months of lockdown (April and May 2020), have any of the following aspects of your life changed**?

Please mark one answer on each line.

If you didn't do the activity before, and aren't doing it now, please select 'not applicable'.

| | **Decreased a lot** | **Decreased a little** | **Stayed the same** | **Increased a little** | **Increased a lot** | **Not applicable** |
|---|---|---|---|---|---|---|
| | ◻ | ◻ | ◻ | ◻ | ◻ | ◻ |

| | | | | | | |
|---|---|---|---|---|---|---|
| Number of home-cooked meals you eat | | | | | | |
| Number of meals you eat in a day | ☐ | ☐ | ☐ | ☐ | ☐ | ☐ |
| Number of snacks you eat in a day | ☐ | ☐ | ☐ | ☐ | ☐ | ☐ |
| Amount of physical activity/exercise you do | ☐ | ☐ | ☐ | ☐ | ☐ | ☐ |
| Amount you sleep | ☐ | ☐ | ☐ | ☐ | ☐ | ☐ |
| Amount you smoke | ☐ | ☐ | ☐ | ☐ | ☐ | ☐ |

**Q45. The following questions are about how you might have been feeling or acting recently. For each statement, please tell us how you have been feeling during the last two weeks.**

| | **Not true** | **Sometimes true** | **True** |
|---|---|---|---|
| I felt miserable or unhappy | ☐ | ☐ | ☐ |
| I didn't enjoy anything at all | ☐ | ☐ | ☐ |
| I felt so tired I just sat around and did | ☐ | ☐ | ☐ |
| I was very restless | ☐ | ☐ | ☐ |
| I felt I was no good anymore | ☐ | ☐ | ☐ |
| I cried a lot | ☐ | ☐ | ☐ |
| I found it hard to think properly or [illegible]rate | ☐ | ☐ | ☐ |
| I hated myself | ☐ | ☐ | ☐ |

| | | | |
|---|---|---|---|
| I was a bad person | ◻ | ◻ | ◻ |
| I felt lonely | ◻ | ◻ | ◻ |
| I thought nobody really loved me | ◻ | ◻ | ◻ |
| I thought I could never be as good as | ◻ | ◻ | ◻ |
| I did everything wrong | ◻ | ◻ | ◻ |

**Q46. Below are some statements about feelings and thoughts. Please select the answer that best describes your experience during the last two weeks.**

| | **None of the time** | **Rarely** | **Some of the time** | **Often** | **All of the time** |
|---|---|---|---|---|---|
| I’ve been feeling optimistic about the future | ◻ | ◻ | ◻ | ◻ | ◻ |
| I've been feeling useful | ◻ | ◻ | ◻ | ◻ | ◻ |
| I've been feeling relaxed | ◻ | ◻ | ◻ | ◻ | ◻ |
| I've been feeling interested in other people | ◻ | ◻ | ◻ | ◻ | ◻ |
| I've had energy to spare | ◻ | ◻ | ◻ | ◻ | ◻ |
| I've been dealing with problems well | ◻ | ◻ | ◻ | ◻ | ◻ |
| I've been thinking clearly | ◻ | ◻ | ◻ | ◻ | ◻ |
| I've been feeling good about myself | ◻ | ◻ | ◻ | ◻ | ◻ |

| | | | | | |
|---|---|---|---|---|---|
| I've been feeling close to other people | ◻ | ◻ | ◻ | ◻ | ◻ |
| I've been feeling confident | ◻ | ◻ | ◻ | ◻ | ◻ |
| I've been able to make up my own mind about things | ◻ | ◻ | ◻ | ◻ | ◻ |
| I've been feeling loved | ◻ | ◻ | ◻ | ◻ | ◻ |
| I've been interested in new things | ◻ | ◻ | ◻ | ◻ | ◻ |
| I've been feeling cheerful | ◻ | ◻ | ◻ | ◻ | ◻ |

**Q47. The following questions are about feelings you may have experienced after the pandemic.**
**Over the last 2 weeks, how often have you been bothered by the following problems?**

| | **Not at all** | **Less than half the days** | **More than half the days** | **Nearly every day** |
|---|---|---|---|---|
| Feeling nervous, anxious or on edge | ◻ | ◻ | ◻ | ◻ |
| Not being able to stop or control worrying | ◻ | ◻ | ◻ | ◻ |
| Worrying too much about different things | ◻ | ◻ | ◻ | ◻ |
| Trouble relaxing | ◻ | ◻ | ◻ | ◻ |
| Being so restless that it is hard to sit still | ◻ | ◻ | ◻ | ◻ |
| Becoming easily annoyed or irritable | ◻ | ◻ | ◻ | ◻ |

| | | | | |
|---|---|---|---|---|
| Feeling afraid as if something awful might happen | ☐ | ☐ | ☐ | ☐ |

**Q48. The following questions are about how you might react to stressful situations. For each question please tell us in the last month how often you have been feeling:**

| | **Never** | **Almost never** | **Sometimes** | **Fairly often** | **Very often** |
|---|---|---|---|---|---|
| Upset because of something that happened unexpectedly | ☐ | ☐ | ☐ | ☐ | ☐ |
| That you were unable to control the important things in your life | ☐ | ☐ | ☐ | ☐ | ☐ |
| Nervous and 'stressed' | ☐ | ☐ | ☐ | ☐ | ☐ |
| Confident about your ability to handle your personal problems | ☐ | ☐ | ☐ | ☐ | ☐ |
| That things were going your way | ☐ | ☐ | ☐ | ☐ | ☐ |
| That you could not cope with all the things that you had to do | ☐ | ☐ | ☐ | ☐ | ☐ |
| That you have been able to control irritations in your life | ☐ | ☐ | ☐ | ☐ | ☐ |
| That you were on top of things | ☐ | ☐ | ☐ | ☐ | ☐ |
| Angered because of things that were outside of your control | ☐ | ☐ | ☐ | ☐ | ☐ |
| That difficulties were piling up so high that you could not overcome them | ☐ | ☐ | ☐ | ☐ | ☐ |

**Q49. Thinking back to January or February 2020, how do you feel you are able to cope with day-to-day life now compared to then?**

- ☐ Much worse
- ☐ A little worse
- ☐ About the same
- ☐ A little better
- ☐ Much better

**Q50. How well would you say you have been managing financially since July 2020?**

- ☐ Living comfortably

- ◻ Doing all right
- ◻ Just about getting by
- ◻ Finding it quite difficult
- ◻ Finding it very difficult

### 4. Interpretation and details of the scales used to measure the psycosocial impacts of COVID-19 pandemic

Short Mood and feeling questionnaire (sMFQ) is a thirteen-item questionnaire derived from a 33 item Mood and Feeling questionnaire that is designed to assess depression symptoms in children and adults. Each item is rated on a 3-point scale i.e., 0 for not true, 1 for sometimes true and 2 for true and the total score ranges from 0 to 26. Score of 12 or higher may indicate the presence of depression.
The Warwick-Edinburgh Mental Well-being Scale (WEMWBS) is used to assess the mental wellbeing of the general population. It is a 14-item scale with 5 response categories based on 5-point Likert scale: 1 for "none of the time", 2 for "rarely", 3 for "some of the time", 4 for "often" and 5 for "all of the time". The overall response is calculated by adding the scores of each item, with scores ranging from 14 to 70 points and higher scores representing a higher level of mental wellbeing.
Generalised Anxiety Disorder Assessment (GAD-7) is a 7-item scale used for the screening of generalized anxiety disorders, rated on a four-point Likert scale: 0 for "not at all", 1 for "less than half the days", 2 for "more than half the days" and 4 for "nearly every day". The sum score ranges from 0 to 21, with three cutoff points 5,10 and 15, and anxiety is classified as 0-4 (none/normal), 5-9 (mild), 10-14 (moderate) and 15-21 (severe).
The Perceived Stress Scale (PSS) is a 10-item stress assessment instrument, that measures the degree of perceived stress based upon situations faced. Each item is scored based on 5-point scale: 0 for "never", 1 for "almost never", 2 for "sometimes", 3 for "fairly often" and 4 for "very often". The total score can range from 0 to 40, which is further categorized as 0-13 (low stress), 14-36 (moderate stress) and 27-40 (high stress).
Changes in different aspects of everyday life, like eating, physical exercise, smoking and sleeping, were accessed by asking simple questions: "Thinking about life now, compared to the early months of lockdown (April and May 2020), have any of the following aspects of your life changed?" and options were given as "decreased a lot", "decreased a little", "stayed the same", "increased a little" and "not applicable". Similarly, the financial status was addressed by asking questions: a) "How well would you say you have been managing financially since March 2020 (since the pandemic)?" with options "doing alright", "just about getting by", "finding it quite difficult" and "finding it very difficult" b) "Thinking back to January or February 2020, how do you feel you are able to cope with day-to-day life now compared to then?" with options "much worse", "a little worse", "about the same", "a little better" and "much better".

### 5. Literature Search strategy

**PubMed**

Advanced search: (((COVID-19) OR (SARS-CoV-2)) OR (long COVID)) AND (Mental health)) OR (psychological impacts)) OR (psychosocial impacts)

My NCBI filters
Species Human
Language English

Date: 2019-30Jan 2023
